# A randomized double-blind placebo-controlled Phase 1 trial of PXS-6302, a topical lysyl oxidase inhibitor, in mature scars

**DOI:** 10.1101/2025.02.12.25321764

**Authors:** Natalie Morellini, Peijun Gong, Suzanne Rea, Helen Douglas, Phuoc Hao Ho, Barry Cense, Brendan F. Kennedy, Wolfgang Jarolimek, Brett Charlton, Alison Findlay, Joanna Leadbetter, Fiona M. Wood, Mark W. Fear

**Affiliations:** Burn Injury Research Unit, School of Biomedical Sciences, The University of Western Australia, Perth, Western Australia, Australia; Fiona Wood Foundation, Fiona Stanley Hospital, Murdoch, Western Australia, Australia; Department of Electrical, Electronic & Computer Engineering, School of Engineering, The University of Western Australia, Perth, Western Australia, Australia; Burns Service Western Australia, WA Department of Health, Fiona Stanley Hospital, Murdoch, Western Australia, Australia; BRITElab, Harry Perkins Institute of Medical Research, QEII Medical Centre, Nedlands and Centre for Medical Research, The University of Western Australia, Perth, Western Australia, Australia; Institute of Physics, Faculty of Physics, Astronomy and Informatics, Nicolaus Copernicus University, Toruń, Poland; Syntara Ltd (formerly Pharmaxis Ltd), Sydney, New South Wales, Australia

## Abstract

Skin scars are a significant clinical challenge, with poor appearance and increased tissue stiffness affecting both physical and psychological wellbeing. Lysyl oxidases are a family of enzymes that catalyze collagen crosslinking, a key factor in scar pathophysiology. Here we report a randomized, double-blind, placebo-controlled Phase 1 clinical trial to assess the safety and tolerability of PXS-6302, a topical pan-lysyl oxidase inhibitor, in treating mature scars (ACTRN12621001545853). Fifty participants with scars were enrolled and PXS-6302 or placebo cream applied to a 10 cm^2^area for three months. No severe adverse events were reported. All treatment-related adverse events (AEs) were localized skin reactions. Treatment with PXS-6302 significantly inhibited lysyl oxidase activity (66%). Hydroxyproline (a marker for collagen) and total protein concentration in the scar were significantly reduced in the PXS-6302 treatment group compared to placebo. Optical coherence tomography (OCT) was used to measure vascularity and attenuation (a marker of extracellular matrix composition). PXS-6302 treatment significantly increased vessel density at 3 months compared to baseline. Tissue attenuation was also significantly increased in PXS-6302 treated participants compared to baseline, suggesting extracellular matrix was becoming increasingly similar to normal skin. No significant differences between placebo and PXS-6302 treatment groups were observed in Patient Observer Scar Assessment Scale (POSAS) scores at study conclusion. To our knowledge, this study represents the first demonstration of a safe and effective pharmaceutical intervention that significantly improves the molecular composition of established scar extracellular matrix in humans. Pan-lysyl oxidase inhibition therefore represents a potential paradigm shift for the amelioration of scarring.

**One Sentence Summary:** Mature scars remodeled after treatment with a topical cream for three months.

## INTRODUCTION

Scar formation following skin trauma is a global health issue with significant physical and psychological impacts. Scar tissue is characterized by excess, densely packed collagen in the dermal extracellular matrix (ECM). This excess and densely packed collagen underpins the poor appearance and increased stiffness of scar tissue (*1–2*). Currently, treatments for significant established scars primarily involve revision surgery, laser treatment or intralesional injections of corticosteroids (*3*). However, these interventions can be painful, invasive, expensive and oftentimes ineffective. It is, therefore, critical that new therapeutic approaches to ameliorate scarring are developed.

Lysyl oxidases catalyze the oxidation of lysine residues in collagen and elastin, resulting in the formation of aldehydes that spontaneously react, either with other aldehydes or with unmodified lysine residues (*4*). This process results in the formation of cross-links that tightly bind collagen strands, increasing mechanical strength and reducing collagen solubility and, ultimately, rendering it more resistant to degradation and turnover (*5*). Under normal physiological conditions, cross-linking plays a vital role in skin homeostasis and ECM maintenance. However, in pathological conditions and diseases, increased production and reduced degradation of ECM favors excessive cross-link formation and results in increased stiffness and fibrosis (*6–9*).

PXS-6302 is a specific and selective inhibitor of all lysyl oxidases (*10*). Upon enzymatic processing, the allylamine portion of the small molecule binds rapidly and irreversibly to the catalytic site, effectively blocking lysyl oxidase activity. Enzymatic activity can only be restored following synthesis of new enzyme. Previously we have shown that PXS-6302 treatment leads to a reduction in collagen cross-linking and collagen deposition *in vitro* and *in vivo*, and improved scar appearance in a porcine model of excision and burn injury when treated in the acute phase (*10*). We hypothesized that in established scar tissue, PXS-6302-mediated inhibition of lysyl oxidases would reduce collagen stability and promote ECM remodeling. To test this, we conducted a randomized, double-blind, placebo-controlled, single site Phase 1c clinical trial (ACTRN12621001545853). The primary objective was to investigate the safety and tolerability of multiple applications of PXS-6302 over a 3-month period to a mature scar (greater than one year from injury). The secondary objectives of the trial included examining the pharmacokinetics (PK) of PXS-6302, as well as assessing the impact of PXS-6302 treatment on the ECM of the scar and scar appearance.

## RESULTS

### Study participants

A total of 50 participants, male and female, aged between 18 and 60 years with established scars (>1-year post-injury) of a minimum 10 cm^2^ surface area were enrolled and evaluated in two cohorts (Fig. 1). Cohort 1 included 8 participants treated daily with 2% PXS-6302 cream (200 µL per 10 cm^2^; 4 mg PXS-6302) over 3 months. Cohort 2 included 42 evaluable participants randomized 1:1 to 2% PXS-6302 or placebo cream application to a 10 cm^2^ area of scar. Cream was applied daily for the first week and 3 times per week thereafter for a total of 3 months. Participants were recruited from 21^st^ January 2022 to 20^th^ December 2022, with the last participant visit conducted on 14^th^ March 2023.

**Fig. 1.**
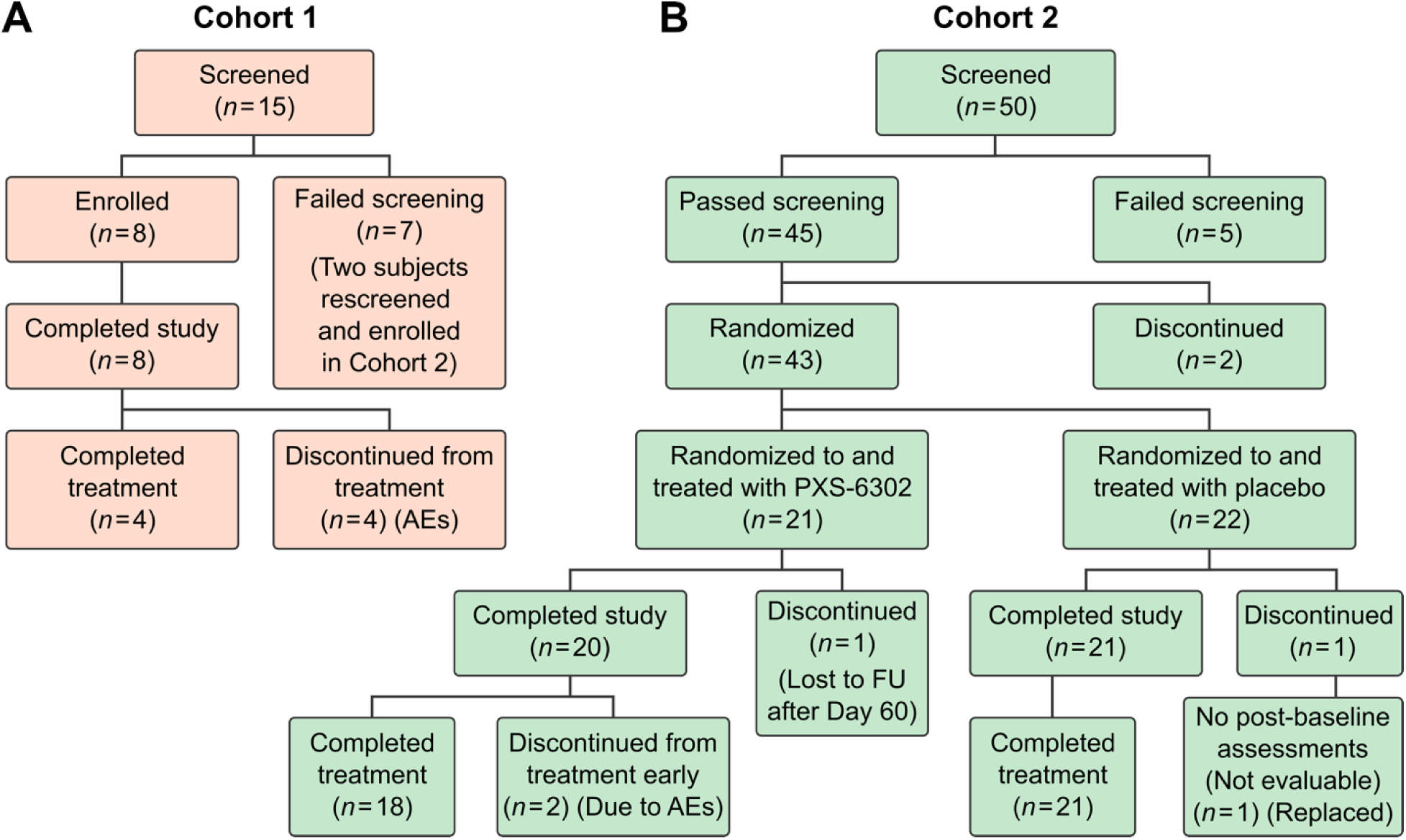
Disposition - Screening and randomization flow chart of Cohort 1 and Cohort 2. (A) Cohort 1 was an open-label single arm study involving 8 participants all receiving PXS-6302 once daily. (B) Cohort 2 was a double-blind randomized study with 1:1 allocation of PXS-6302:placebo with 21 evaluable patients in each arm and 18 completing PXS-6302 treatment. AE: adverse event; FU: follow-up.

For Cohort 1, a total of 15 participants were assessed for eligibility, 8 were enrolled and all received PXS-6302. For Cohort 2, a total of 50 participants (including 2 who were ineligible for Cohort 1) were assessed for eligibility, 45 were enrolled, 2 withdrew prior to randomization and 43 were confirmed for randomization; 21 were randomized to PXS-6302 and 22 to placebo. Two participants were lost to follow-up; one from the placebo group was not evaluable as they did not return after baseline and was thus replaced; one from the PXS-6302 group did not return after the Day 60 visit and was not replaced.

Baseline characteristics of the enrolled patients are shown in Table 1. In Cohort 1, all scars were burn scars, mostly from severe burn injuries, and the median time since injury was 4.0 (range 2.2 to 13) years. In Cohort 2, 66.7% of participants had burn scars from non-severe injuries and the median time since injury was 4.14 (range 1 to 54) years. In Cohort 2, there were notable baseline imbalances in the aetiology of scar between placebo (57.1% burns) and PXS-6302 (76.2% burns) treatment groups, as well as the time since injury, with placebo group having a greater number of participants with scars under 2 years old (33.3% ≤2 years) compared to PXS-6302 treatment participants (14.3% ≤2 years).

**Table 1.**
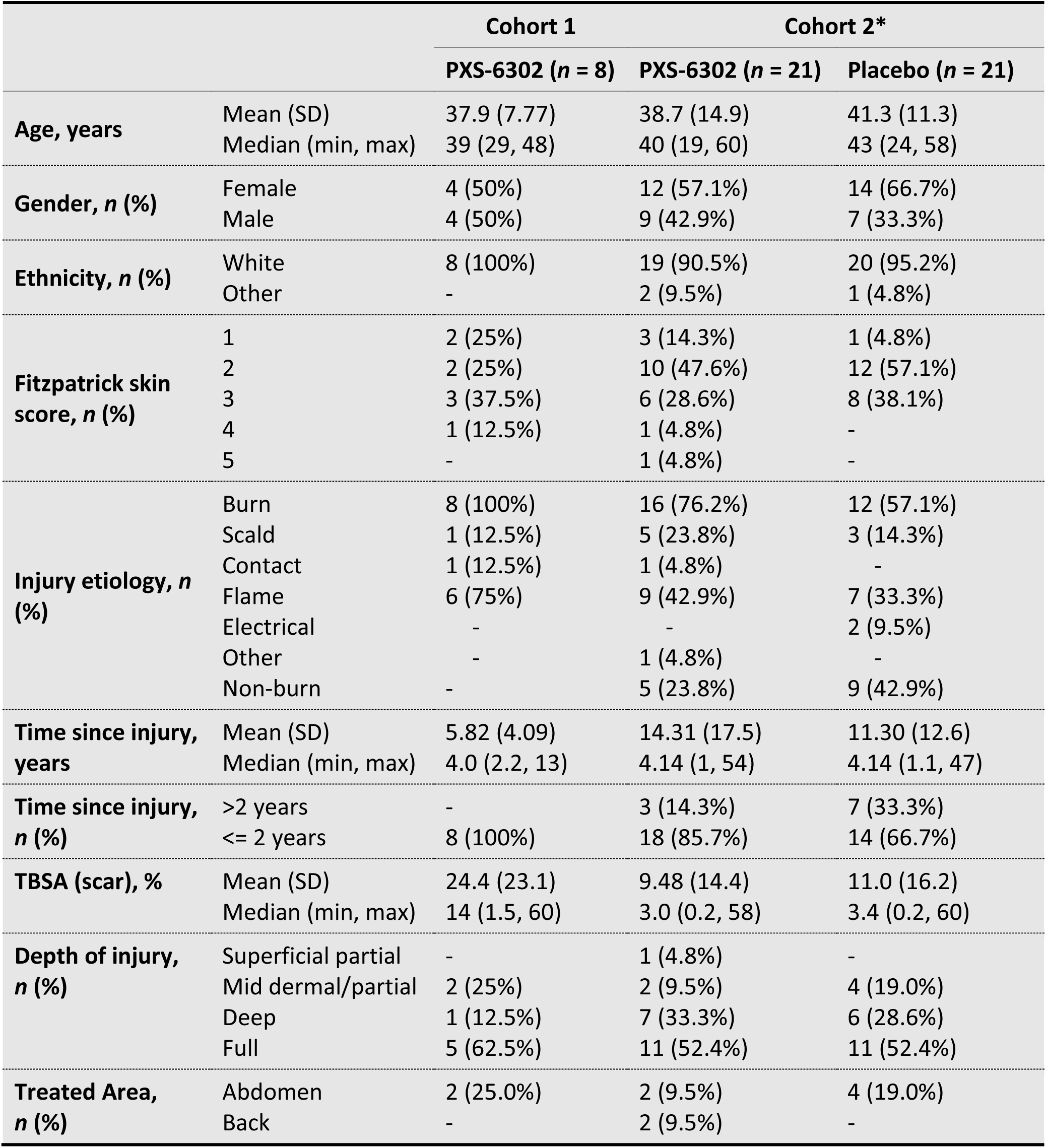
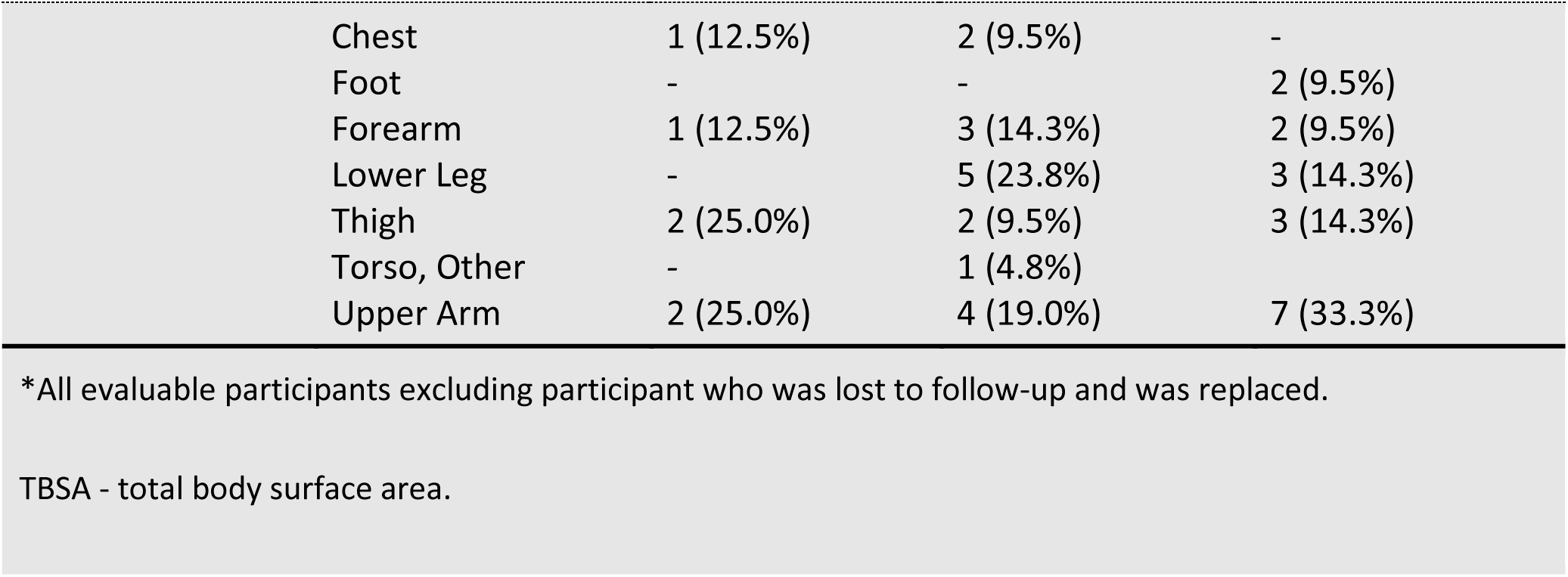
Demography and baseline characteristics.

| Table 1. Demography and baseline characteristics. |  |  |  |  |
| --- | --- | --- | --- | --- |
|  |  | Cohort 1 | Cohort 2* |  |
|  |  | PXS-6302 (n = 8) | PXS-6302 (n = 21) | Placebo (n = 21) |
| Age, years | Mean (SD) | 37.9 (7.77) | 38.7 (14.9) | 41.3 (11.3) |
|  | Median (min, max) | 39 (29, 48) | 40 (19, 60) | 43 (24, 58) |
| Gender, n (%) | Female | 4 (50%) | 12 (57.1%) | 14 (66.7%) |
|  | Male | 4 (50%) | 9 (42.9%) | 7 (33.3%) |
| Ethnicity, n (%) | White | 8 (100%) | 19 (90.5%) | 20 (95.2%) |
|  | Other | - | 2 (9.5%) | 1 (4.8%) |
| Fitzpatrick skin score, n (%) | 1 | 2 (25%) | 3 (14.3%) | 1 (4.8%) |
|  | 2 | 2 (25%) | 10 (47.6%) | 12 (57.1%) |
|  | 3 | 3 (37.5%) | 6 (28.6%) | 8 (38.1%) |
|  | 4 | 1 (12.5%) | 1 (4.8%) | - |
|  | 5 | - | 1 (4.8%) | - |
| Injury etiology, n (%) | Burn | 8 (100%) | 16 (76.2%) | 12 (57.1%) |
|  | Scald | 1 (12.5%) | 5 (23.8%) | 3 (14.3%) |
|  | Contact | 1 (12.5%) | 1 (4.8%) | - |
|  | Flame | 6 (75%) | 9 (42.9%) | 7 (33.3%) |
|  | Electrical | - | - | 2 (9.5%) |
|  | Other | - | 1 (4.8%) | - |
|  | Non-burn | - | 5 (23.8%) | 9 (42.9%) |
| Time since injury, years | Mean (SD) | 5.82 (4.09) | 14.31 (17.5) | 11.30 (12.6) |
|  | Median (min, max) | 4.0 (2.2, 13) | 4.14 (1, 54) | 4.14 (1.1, 47) |
| Time since injury, n (%) | >2 years | - | 3 (14.3%) | 7 (33.3%) |
| | ≤ 2 years | 8 (100%) | 18 (85.7%) | 14 (66.7%) |
| TBSA (scar), % | Mean (SD) | 24.4 (23.1) | 9.48 (14.4) | 11.0 (16.2) |
|  | Median (min, max) | 14 (1.5, 60) | 3.0 (0.2, 58) | 3.4 (0.2, 60) |
| Depth of injury, n (%) | Superficial partial | - | 1 (4.8%) | - |
|  | Mid dermal/partial | 2 (25%) | 2 (9.5%) | 4 (19.0%) |
|  | Deep | 1 (12.5%) | 7 (33.3%) | 6 (28.6%) |
|  | Full | 5 (62.5%) | 11 (52.4%) | 11 (52.4%) |
| Treated Area, n (%) | Abdomen | 2 (25.0%) | 2 (9.5%) | 4 (19.0%) |
|  | Back | - | 2 (9.5%) | - |

|  |  |  |  |
| --- | --- | --- | --- |
| Chest | 1 (12.5%) | 2 (9.5%) | - |
| Foot | - | - | 2 (9.5%) |
| Forearm | 1 (12.5%) | 3 (14.3%) | 2 (9.5%) |
| Lower Leg | - | 5 (23.8%) | 3 (14.3%) |
| Thigh | 2 (25.0%) | 2 (9.5%) | 3 (14.3%) |
| Torso, Other | - | 1 (4.8%) |  |
| Upper Arm | 2 (25.0%) | 4 (19.0%) | 7 (33.3%) |

\*All evaluable participants excluding participant who was lost to follow-up and was replaced.
TBSA - total body surface area.

### Safety profile of PXS-6302

A summary of all Adverse Events (AEs) considered related to the study drug is documented in Table 2. In Cohort 1, AEs considered related to study drug by investigators were reported in 4 out of 8 participants, with first events occurring between Day 14 and Day 36. These AEs were application site reactions, where 3 were graded as mild and 1 as moderate. The 3 mild reactions involved pruritis and erythema at the application site only, whereas the moderate reaction involved spreading of erythema to other areas. All participants recovered within 1-2 weeks of ceasing PXS-6302 and did not require any other treatment or intervention. Once recovered, 3 out of the 4 participants applied the cream again. However, after one or two applications, they all recorded another application site reaction, and therefore permanently ceased treatment. Despite discontinuing treatment early, all participants who had an adverse reaction completed the study and attended all their appointments. As a result of the adverse site reactions in Cohort 1, the dosing frequency for Cohort 2 was reduced to daily application for the first 7 days followed by three times per week until the end of the study.

**Table 2.** All treatment related adverse events observed during both Cohort 1 and Cohort 2 studies. No serious adverse events were observed. Adverse events related to the treatment were all localized skin reactions.

| Cohort | Treatment | Adverse event | Severity | Onset Day | Action with study treatment | Outcome |
| --- | --- | --- | --- | --- | --- | --- |
| 1 | PXS-6302 | Application site reaction (erythema/pruritis) | Mild | 14 | Ceased treatment | Recovered |
|  | PXS-6302 | Application site reaction (erythema/pruritis) | Moderate (and then Mild) | 36 and 58 | Ceased treatment until recovered. Re-started on Day 58. Treatment permanently stopped when event re-occurred. | Recovered |
|  | PXS-6302 | Application site reaction (erythema/pruritis) | Mild | 28 and 55 | Ceased treatment until recovered. Restarted on Day 53. Treatment permanently stopped when event re-occurred. | Recovered |
|  | PXS-6302 | Application site reaction (erythema/pruritis) | Mild | 25 and 39 | Ceased treatment until recovered. Restarted on Day 39. Treatment permanently stopped when event re-occurred. | Recovered |
| 2 | Placebo | Application site erythema | Mild | 26 | Stopped treatment for a week and resumed when resolved on Day 33 | Recovered |
|  | PXS-6302 | Application site paresthesia | Mild | 15 | Continued treatment-monitor | Recovered |
|  | PXS-6302 | Application site erythema | Mild | 85 | Study treatment already ceased-end of study | Recovered |
|  | PXS-6302 | Application site reaction (erythema/pruritis) | Mild | 1 | Continued treatment-monitor | Recovered |
|  | PXS-6302 | Application site reaction (erythema/pruritis) spread to adjacent site | Moderate | 52 | Ceased treatment | Recovered |
|  | PXS-6302 | Application site erythema | Mild | 63 | Ceased treatment | Recovered |

In Cohort 2, AEs considered related to the study drug by investigators were reported in 5 participants treated with PXS-6302 and in 1 participant treated with placebo. In the placebo group, one participant experienced mild erythema at the application site on Day 26. As a precautionary measure, the treatment was temporarily discontinued for one week. Subsequently, the participant resumed treatment without experiencing any further AEs. In the PXS-6302 group, four AEs were graded as mild, and one was graded as moderate. The mild AEs involved a reaction at the application site only, while the moderate AE spread to another area that was in contact with the application site. Two participants reported mild application site paraesthesia and/or pruritis during the first 2 weeks of application which resolved by itself by the Day 28 appointment. Mild application site erythema was observed in one participant on the final Day 90 appointment; however this did not impact treatment. Mild application site erythema was also observed in another participant on Day 63 who ceased treatment as a result. The AE was resolved by Day 73. One participant experienced a moderate AE at Day 52 of application site erythema and pruritis which spread to an adjacent area that was in contact and discontinued treatment as a result. This resolved completely at Day 67. Both participants who discontinued treatment early completed the study and attended the final Day 90 appointment. There were no clinically significant changes observed at any time-point in clinical chemistry, hematology or vital signs measured.

### Pharmacodynamics and pharmacokinetics

In Cohort 1, PXS-6302 tissue concentration and lysyl oxidase activity were measured in scar biopsies on Days 7, 28 and 90. The mean (±SD) drug concentrations in scar biopsies were similar at Days 7 (179314 ± 160317 ng/g) and 28 (103349 ± 82222 ng/g) suggesting steady state was achieved (Fig. 2A). PXS-6302 levels in the blood plasma were measured at Days 7, 21, 28, 60 and 90 with very low or non-measurable levels (lower limit of quantification 0.01 ng/mL) detected at all time-points (Fig. 2B). Furthermore, scar biopsies showed a significant reduction in lysyl oxidase activity (74.4 ± 22.8% inhibition, p<0.001) at Day 7 that was also observed at Day 28 (83.6 ± 11.6%, p<0.001) when compared to baseline (Fig. 2C). At Day 90, in three participants who completed treatment, lysyl oxidase inhibition was 75.9 ± 12% when measured within 3 days of the last dose (one additional patient reached day 90, excluded from mean calculation due to outlying value; this patient reported missing multiple doses (Fig. 2C))

**Fig. 2.**
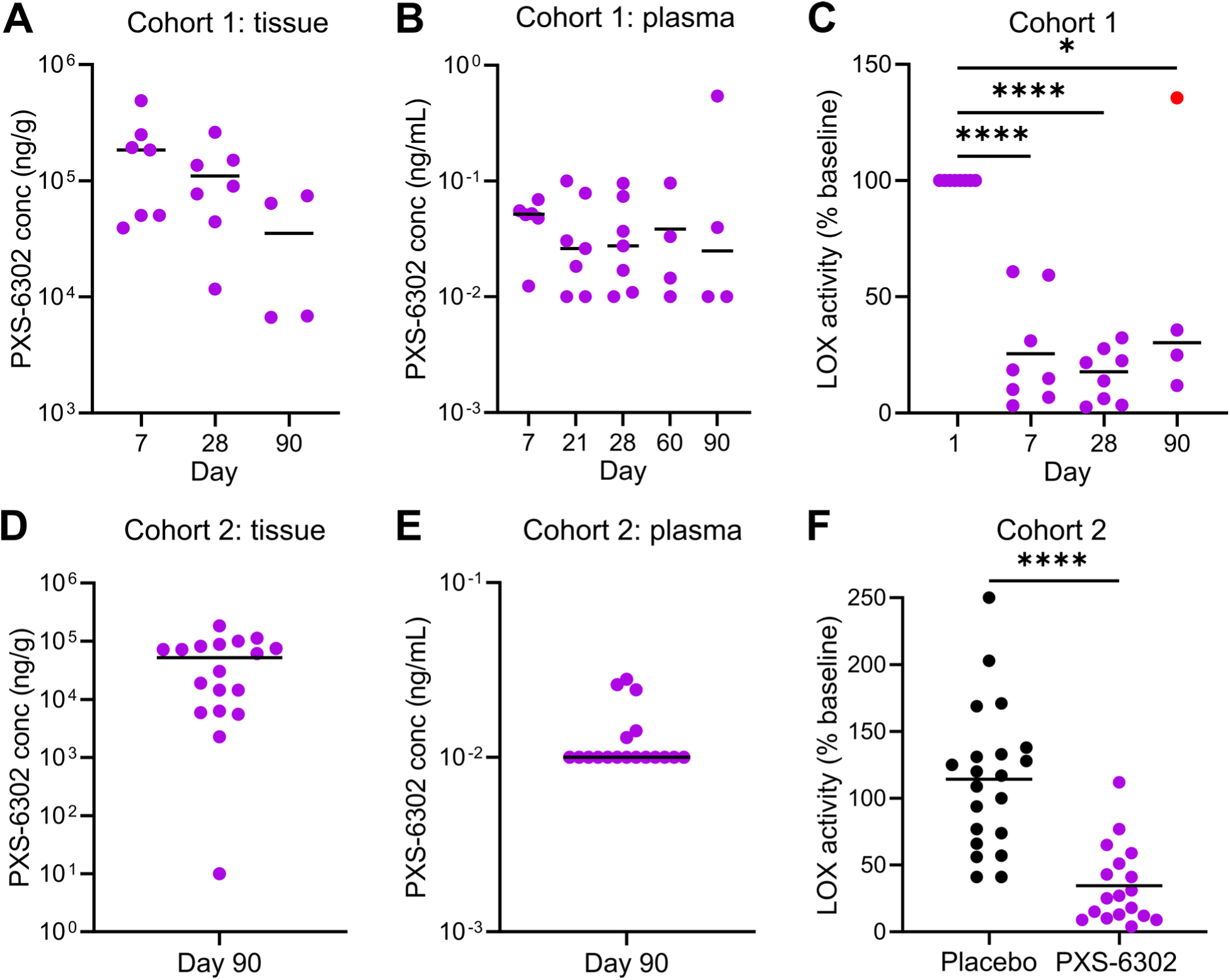
Topical application of PXS-6302 results in effective local inhibition of Lysyl oxidase activity. **(A)** Concentration of PXS-6302 in tissue in Cohort 1. **(B)** PXS-6302 concentration in plasma for Cohort 1. **(C)** Lysyl oxidase activity as percent of baseline (day 1) activity in cohort 1. **(D)** Concentration of PXS-6302 in tissue in Cohort 1. **(E)** PXS-6302 concentration in plasma for Cohort 1. **(F)** Lysyl oxidase activity as percent of baseline (day 1) activity in cohort 2 for placebo and PXS0-6302 treated groups at Day 90. Graphs are presented as concentrations (PXS-6302), percentage of baseline lLysyl oxidase activity), or percentage change from baseline, normalized against biopsy weight (other measures). 10^-2^ ng/mL is the lower limit of detection in plasma which was reported when the measured levels were below the threshold of quantitation. Horizontal line: mean (excluding outlier where applicable). Average of biopsy weight was identical between placebo and PXS-6302 cohorts. Cohort 2 data shows all participants that completed treatment (*n* = 18 PXS-6302 group, *n* = 21 placebo group). Red point indicates an outlier. **** p<0.0001, * p<0.05.

In Cohort 2, drug concentration in scar biopsies and blood plasma were measured up to 72 hours after the final drug application. PXS-6302 concentrations were high in the scar (52568 ± 50314 ng/g, Fig. 2D) and very low/negligible in plasma (0.013 ± 0.006 ng/mL, Fig. 2E (n=18 completed participants)), indicating that PXS-6302 was absorbed and remained present in the scar tissue after repeated dosing. At Day 90 PXS-6302 significantly reduced lysyl oxidase activity compared to baseline (66 ± 29% inhibition, p<0.0001, in all participants that completed treatment) and placebo (mean difference between groups ± SEM, 79.8 ± 14.2%, p<0.0001, (Fig. 2F)), with no significant change compared to baseline in the placebo group (Fig. 2F).

### Collagen and total protein levels in scar tissue

Hydroxyproline (Hyp) and total protein concentration were measured in Cohort 2 to investigate changes in the ECM after 3 months of PXS-6302 treatment. Hyp is a commonly used surrogate for collagen as this amino acid is rare in the majority of proteins but accounts for approximately 10% of the residues in collagen 1. Hyp, when normalized for biopsy weight, was significantly reduced at Day 90 compared to baseline in the PXS-6302 treatment group (80269 ± 20201 ng/mg tissue at baseline and 69142 ± 29563 ng/mg tissue at Day 90, p<0.05; Fig. 3A), with a significant increase observed in the placebo group (65852 ± 16280 ng/mg tissue at baseline and 76433 ± 14244 at Day 90, p<0.01, Fig. 3A). When normalized to biopsy weight, PXS-6302 treatment resulted in an average reduction of 16.3% ± 31.2% (Fig. 3B) in hydroxyproline by Day 90 compared to baseline, with the placebo group exhibiting an increase at Day 90 compared to baseline (21.8% ± 35.3%, Fig. 3B). Compared to placebo, the PXS-6302 group at Day 90 had a significant reduction in Hyp/biopsy weight (−38.1% ±10.8%, p<0.001, Fig. 3B).

**Fig. 3.**
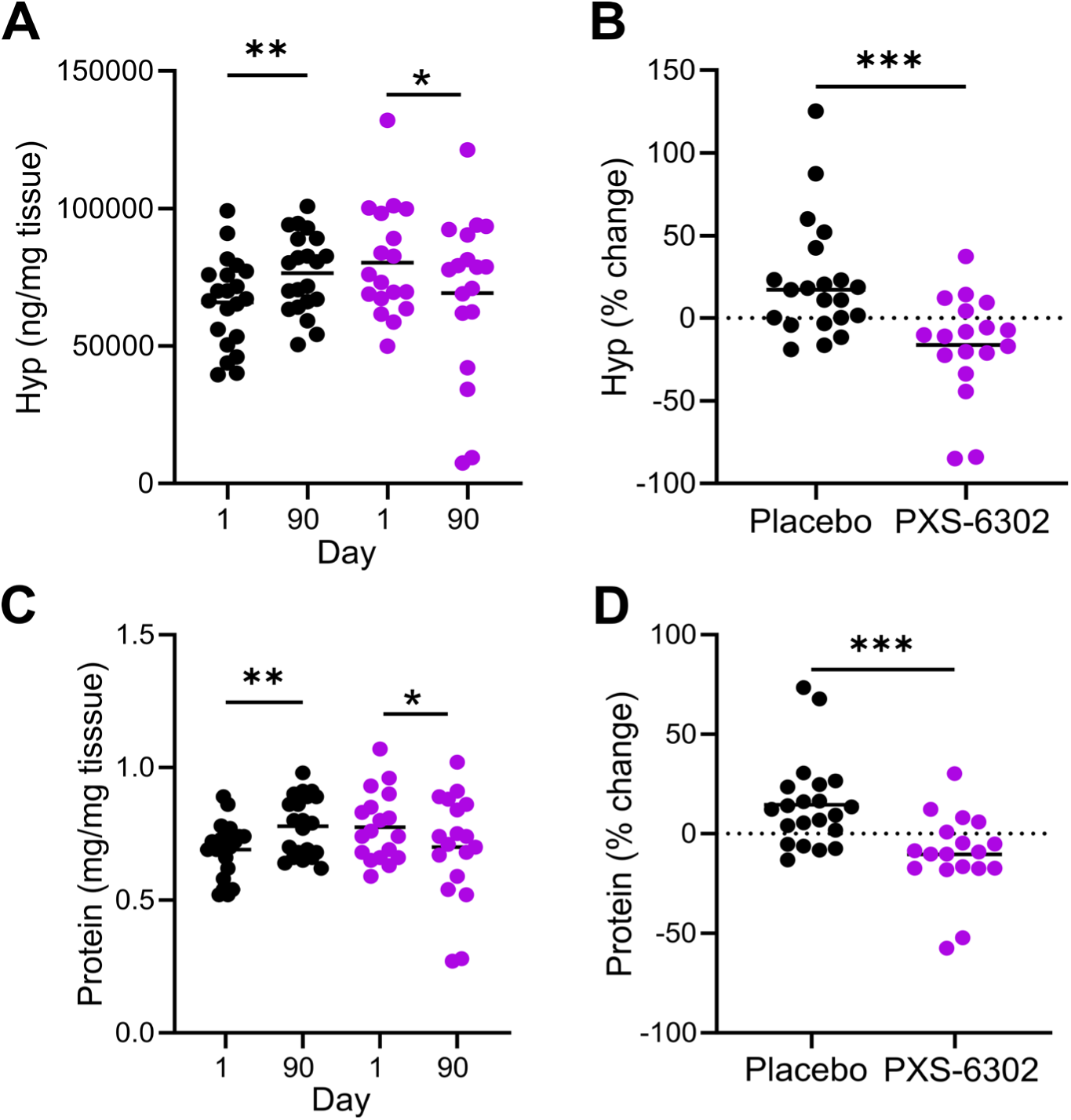
PXS-6302 application significantly reduces both collagen and protein concentration in scar tissue. **(A)** Hyp/tissue weight at baseline and Day 90 **(B)** Change in Hyp at day 90 shown as percent change from baseline **(C)** Protein per mg tissue at day 1 and day 90 **(D)** Protein/tissue weight at day 90 in cohort 2 shown as percent change from baseline (*n* = 18 PXS-6302 group, *n* = 21 placebo group). Horizontal line: mean. P values calculated using Mann-Whitney test (independent groups) or t-test (paired data). *** p<0.001, ** p<0.01, * p<0.05

Total protein when normalized to biopsy weight was also significantly decreased by Day 90 in the PXS-6302 treated group (0.77 mg/mg ± 0.13 tissue at baseline and 0.70 mg/mg ± 0.2 at Day 90, p<0.05, Fig. 3C) with a significant increase in the placebo group at Day 90 (0.69 mg/mg tissue ± 0.11 at baseline and 0.78 mg/mg tissue ± 0.11 at Day 90, p<0.01, Fig. 3C). When normalized to biopsy weight, total protein was reduced by an average of 10.4% ± 20.5% in the PXS-6302 group at Day 90 compared to baseline, whilst the placebo group had a significant increase in total protein at Day 90 of 14.6% ± 12.2% compared to baseline. The PXS-6302 treated group showed a significant reduction in protein at Day 90 of 25.0% ± 6.91% (p<0.001) when compared to placebo (Fig. 3D).

### Optical coherence tomography assessment of vessel density and optical attenuation

To assess the microvasculature and structural changes of the scars, optical coherence tomography (OCT) imaging with a custom data processing method (Fig. 4A) was performed *in vivo* on a subset of 14 consenting participants in Cohort 2 at Day 1 and 90. After unblinding, 7 of these participants were in the PXS-6302 group and 7 in placebo. One participant in the PXS-6302 group was not analyzed due to movement during image acquisition preventing accurate data processing. The projection images of the vessels (skin surface to a depth of 500 µm) at Day 90 as compared to Day 1 baseline showed a significant increase in vessel density (Fig. 4B) for the PXS-6302 treatment group at Day 90 (mean ± standard deviation of the increase: 11.5 ± 5.1%, p=0.031, Fig. 4C) as compared to Day 1, while there was no significant change in the placebo group over time (−1.5 ± 2.4%, Figs. 4C).

**Fig. 4.**
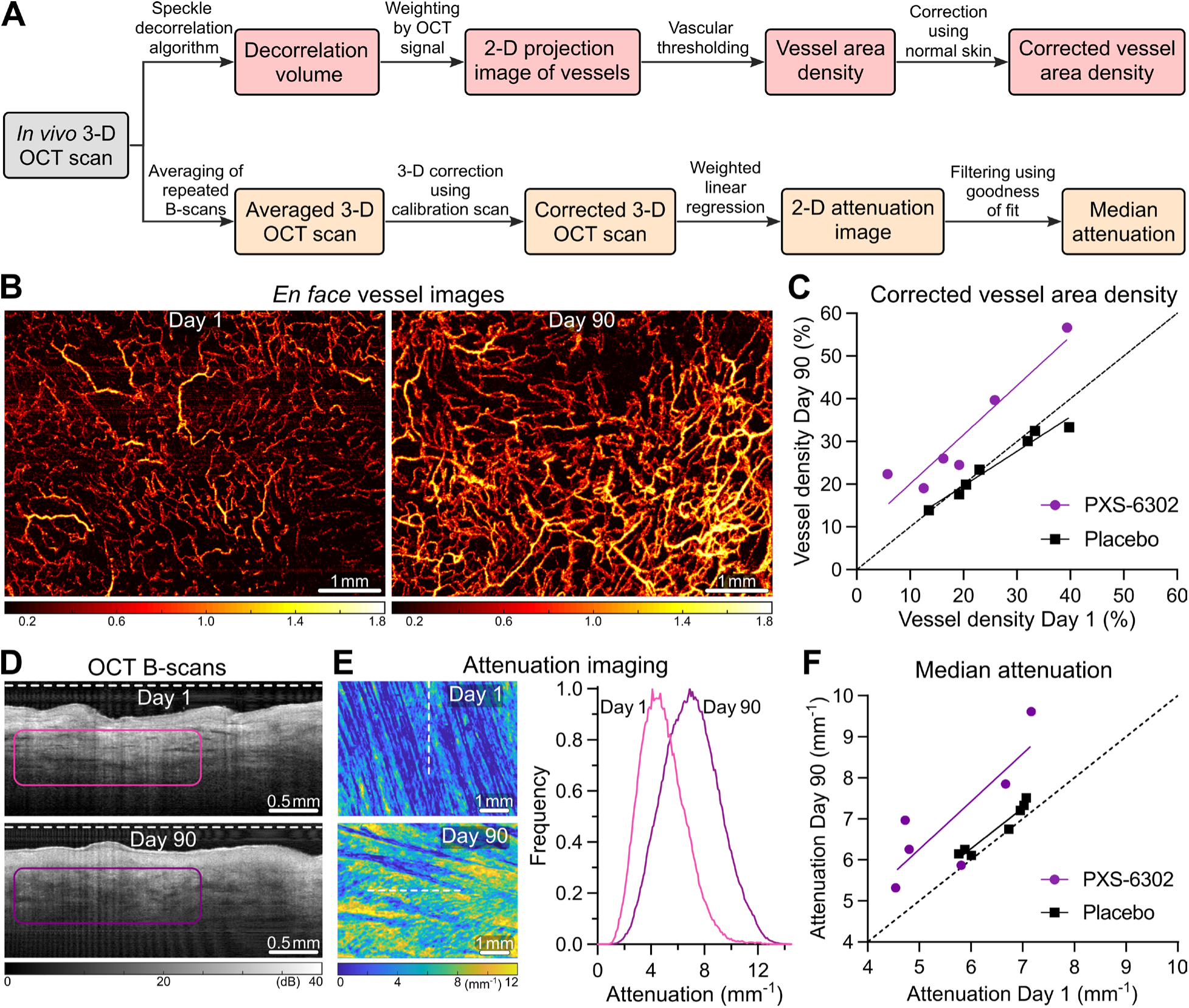
*In vivo* OCT imaging indicates increased vessel area density and optical attenuation of participants treated with PXS-6302. **(A)** The custom data processing method used to process 3-D OCT scans to image and quantify the vasculature and optical attenuation. **(B)** *En face* vessel images from a PXS-6302-treated participant show more abundant vessels after treatment. **(C)** Quantified vessel density shows a significant increase at Day 90 in the PXS-6302 treatment group, with no significant change in the placebo group. **(D)** OCT cross-sectional images from a PXS-6302-treated participant indicate higher optical attenuation along depth at Day 90. (E) *en face* attenuation images at Day 1 and 90 (left) and attenuation distributions (right). **(F)** Measured optical attenuation compared to baseline for both placebo and PXS-6302 treatment groups. Dashed lines in (C) and (F) indicate equal values for Day 1 and 90 (*n* = 6 PXS-6302 group, *n* = 7 placebo group).

OCT images of the scar tissue (Fig. 4D) were used to quantify structural changes in the ECM by measuring changes in the optical attenuation of the tissue. Previously we have shown normal skin has significantly higher attenuation than scar tissue, likely reflecting differences in structure and hydration (*11*). This analysis showed significantly increased attenuation in the PXS-6302 treatment group at Day 90 (Fig. 4E) as compared to Day 1 (1.36 ± 0.9 mm^-1^ (mean ± SD of the increase), p=0.03 (Fig 4F)), while the placebo group showed a significant but much smaller increase over time (0.27 ± 0.16 mm^-1^, p=0.016 (Fig. 4F)). The increase observed in the PXS-6302 treatment group was significantly greater than the increase observed in the placebo group (p=0.03).

### Patient-reported outcome measures

To assess if PXS-6302 treatment improved scar appearance, the Patient and Observer Scar Assessment Scale (POSAS) Version 2.0 was used. POSAS is a subjective tool used to report both patient and observer/clinician observations of the scar (*12*). In the PXS-6302 treated group of Cohort 2, initial baseline patient POSAS scores were higher than those in the placebo group (28.44 ± 8.71 vs 22.95 ± 9.09) as shown in Table 3.

**Table 3.** Patient POSAS scores for Cohort 2 for PXS-6302 and placebo groups at baseline and end of treatment, including individual category scores (Mean (±SD))

| Patient report |  | Pain | Itch | Color | Stiffness | Thickness | Irregularity | Total | Overall |
| --- | --- | --- | --- | --- | --- | --- | --- | --- | --- |
| Placebo<br>(n = 21) | Baseline | 1.10<br>(0.44) | 1.10<br>(0.44) | 5.29<br>(2.37) | 4.38<br>(3.06) | 4.81<br>(2.68) | 6.29<br>(2.70) | 23.0<br>(9.09) | 7.05<br>(2.75) |
|  | Day 90 | 1.00<br>(0.00) | 1.05<br>(0.22) | 4.86<br>(1.71) | 4.29<br>(2.00) | 4.24<br>(2.02) | 5.19<br>(2.48) | 20.6<br>(7.00) | 5.43<br>(2.09) |
|  | Change | -0.10<br>(0.44) | -0.05<br>(0.50) | -0.43<br>(1.75) | -0.10<br>(2.91) | -0.57<br>(2.71) | -1.10<br>(2.57) | -2.33<br>(-7.67) | -1.62<br>(2.38) |
| PXS-6302<br>(n = 18) | Baseline | 1.22<br>(0.94) | 1.61<br>(1.69) | 6.28<br>(2.42) | 5.94<br>(2.69) | 5.83<br>(3.05) | 7.56<br>(2.38) | 28.4<br>(8.71) | 8.22<br>(2.26) |
|  | Day 90 | 1.06<br>(0.24) | 1.44<br>(0.92) | 5.72<br>(2.85) | 4.39<br>(2.79) | 5.06<br>(3.11) | 6.56<br>(2.97) | 24.2<br>(11.41) | 6.56<br>(3.05) |
|  | Change | -0.17<br>(0.99) | -0.17<br>(1.10) | -0.56<br>(2.83) | -1.56<br>(2.36) | -0.78<br>(2.13) | -1.00<br>(2.43) | -4.22<br>(7.36) | -1.67<br>(1.78) |

At all time-points after treatment initiation, patient reported scores were significantly improved when compared to baseline in the PXS-6302 treatment group (Day 28: p=0.021; Day 60: p=0.027; and Day 90: p=0.034 (Figs. 5A, B)), with the significant reduction in score occurring by the 28-day time-point (first time-point measured). No significant difference compared to baseline was observed in patient scores for the placebo group at any time-point (Figs. 5A, B). Surprisingly, the observer scores showed a significant reduction at all time-points in the placebo group (Day 28 p=0.0168, Day 60 p=0.0129 and Day 90 p=0.003 (Figs. 5C, D)), with the largest improvement occurring between months 2 and 3 of treatment, with a significant reduction only observed at Day 28 in the PXS-6302 group (p=0.0288 (Fig. 5C, D)). When corrected for imbalances in baseline POSAS scores, age of scar and scar type, there were no significant differences between placebo and PXS-6302 groups at Day 90.

**Fig. 5.**
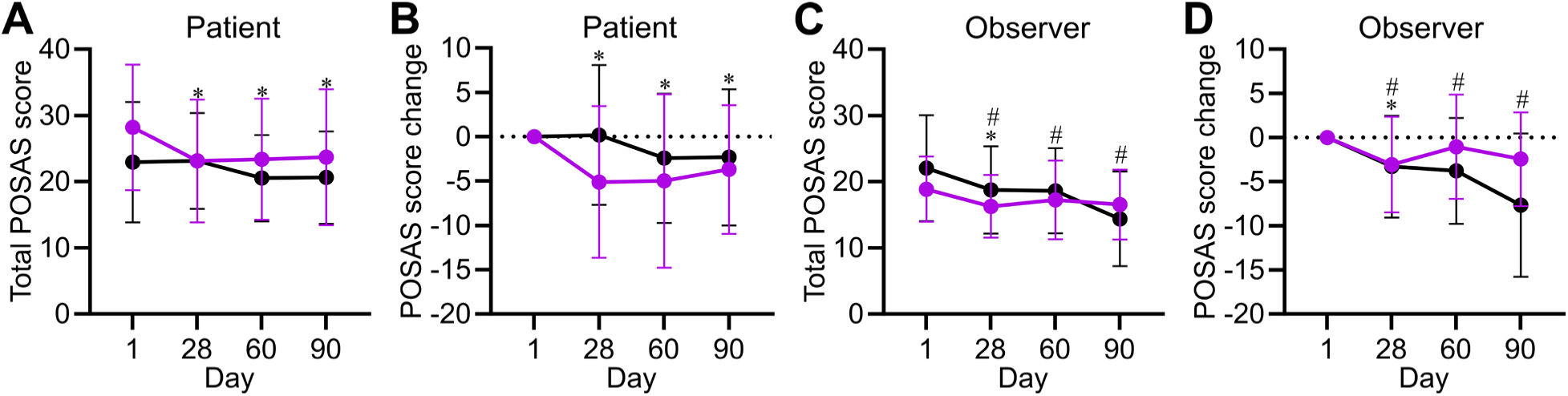
Changes in POSAS scores in PXS-6302 and placebo groups in Cohort 2. **(A)** Patient reported score (total score) over time **(B)**, Change in patient reported score (total score) over time **(C)** Observer reported score (total score) over time **(D)**. Change in observer reported score (total score) over time. Graphs show mean with standard deviation (black line: placebo; purple line: PXS-6302). Dotted horizontal line shows no change. * p<0.05, # p<0.05 for PXS-6302 and placebo respectively when compared to baseline values using all completed patients (n=18 PXS-6302, n=21 placebo).

## DISCUSSION

Herein we report the results of a double-blind placebo-controlled randomized Phase 1 clinical trial examining the safety of the topical pan-lysyl oxidase inhibitor, PXS-6302, when applied to mature scars. We demonstrated that topical application of PXS-6302 three times per week over a period of 3 months was safe, well tolerated and caused a significant improvement in the molecular and structural composition of scar ECM.

Building upon our previous studies in which topical application of PXS-6302 inhibited lysyl oxidases and reduced collagen content in murine and porcine models of scarring (*10*), the present clinical study confirms that PXS-6302 was absorbed into the scar and inhibited lysyl oxidase activity following dosing 3 times a week. When measured up to 72 hours after the last dose, >60% enzyme activity remained inhibited. Lysyl oxidase inhibition appears to trigger a fundamental change to the composition of mature scar tissue, resulting in a subsequent reduction in both collagen and protein content in mature scars. Total protein concentration showed a small but significant increase from baseline to Day 90 in the placebo treated group that was unexpected. This is potentially due to high concentrations of detergents in the biopsies after placebo treatment which soften the skin and solubilize proteins for more effective quantification.

*In vivo* OCT imaging provides a promising non-invasive method to assess microvasculature and extracellular matrix of skin and scar longitudinally (*11, 13*). The results showed increased vessel density and attenuation of the scar tissue at Day 90 for the PXS-6302 treated group. The observed increased microvasculature density in the PXS-6302 group likely is a result of changes in the ECM, predominantly a reduction in collagen content and density, caused by the PXS-6302 treatment. Fibrotic ECM in tumors and in idiopathic lung fibrosis has been shown to restrict tissue perfusion and reduce vessel density (*14–15*), whilst treatment with the anti-fibrotic drug pirfenidone led to increased vessel diameter and perfusion in orthotopic mammary tumor studies (*16*). It has also been previously shown that pan-lysyl oxidase inhibition reduces density of fibrotic tissue, as measured by increased drug concentrations and reduced hydrostatic pressure in fibrotic tissue (*17*) as well as reduced tissue stiffness (*18*). These findings support the hypothesis that the changes observed here in vessel density and attenuation after scar treatment with PXS-6302 reflect structural changes in the ECM. The reduction in optical attenuation of scar tissue compared to matched skin from healthy individuals is largely caused by collagen, hydration and appendages in the skin (*11*). The much greater increase in attenuation observed in the PXS-6302 treatment group suggests a remodeling of tissue in response to PXS-6302 treatment is leading towards a more ‘normal skin’ type ECM with similar values obtained for normal skin in a previous study as seen in the Day 90 PXS-6302 treated scar tissue here (*11*). A future study is still needed to confirm the association of changes in attenuation with structural remodeling of the ECM.

As this was a Phase 1 trial, the design was not optimized for assessing efficacy with respect to scar improvement. Indeed, the inclusion and exclusion scar criteria were deliberately broad to facilitate recruitment focused on demonstrating safety and identifying adverse events. This resulted in many participants having normotrophic, flat and pliable scars at baseline, as well as significant differences in the baseline characteristics of the placebo and PXS-6302 treatment groups that may limit the ability to observe scar improvements as a result of PXS-6302 treatment. Patient and observer POSAS scores were conflicting, highlighting the challenges associated with using subjective scar scales to provide accurate clinical assessments of scar, and the continued need for objective measures, such as OCT, to assist in the development of therapeutics to ameliorate scarring. In contrast, the objective measures applied in this study strongly suggest lysyl oxidase inhibition is an effective mechanism by which to improve scars and warrants a phase 2 efficacy study.

This trial provides encouraging results that safe and effective inhibition of lysyl oxidases can be achieved with a topical therapy, leading to changes in scar ECM. A Phase 2 study will require recruitment of participants with hypertrophic scars, restricted to those with a high POSAS score to better ascertain if the molecular changes that result from lysyl oxidase inhibition improves patient perception and appearance of their scars.

## MATERIALS AND METHODS

### Study design

This study was a Phase 1, randomized, double-blind, placebo-controlled, single center study, enrolling 50 patients with mature scars (≥1-year old). This study was conducted in accordance with the NHMRC statement on ethical conduct in human research (2007) and was approved by the Human Research Ethics Committees at South Metropolitan Health Services and conducted at Fiona Stanley Hospital (RGS0000004980). All eligible participants were provided with a patient information sheet, verbal explanation of the study and written consent was obtained prior to participation. The trial is registered at the Australian And New Zealand Clinical Trials Registry (www.anzctr.org.au/) ACTRN12621001545853.

This study was designed to investigate the safety and tolerability of the lysyl oxidase inhibitor, PXS-6302, formulated in a cream which was applied topically to a 10 cm^2^ area of an established scar for 3 months. The study included two cohorts. An initial cohort (Cohort 1) consisted of 8 participants, who were enrolled in an open label study and treated with 4 mg (200 µL of 2%) PXS-6302 daily. These participants were monitored for systemic levels of PXS-6302 and adverse events for a minimum of 4 weeks prior to enrolling a second cohort (Cohort 2) that was a double-blind 1:1 randomized placebo-controlled trial of PXS-6302 for 21 participants and placebo treatment for 21 participants. The sample size was selected to provide adequate information around the safety and tolerability of multiple applications of PXS-6302 to allow further development to proceed whilst also providing sufficient sample size for a preliminary evaluation of pharmacodynamic and clinical outcomes. The dose of PXS-6302 selected for Cohort 2 was unchanged at 4 mg (2%) but the dosing frequency was modified to be daily dosing for the first week and 3 times per week thereafter.

### Changes in study protocol

The study protocol was amended on three occasions following recruitment of the first participant. The first revision included a change in the inclusion criteria from people with a scar from 1-5 years old to a scar over 1-year old with no maximum time since injury. This change was made to support adequate recruitment for the safety study. Due to the adverse events observed in Cohort 1, a further amendment was added to the protocol to change dosing frequency in Cohort 2 from daily dosing to daily for one week followed by three times per week for the remainder of the study. The final revision included allowing greater investigator discretion in reviewing clinical blood results with respect to exclusion criteria to ensure that healthy participants were not being excluded unnecessarily.

### Participants

As well as recruitment of existing patients of The Western Australian Burns Service, additional patients were recruited through print and media advertising. After providing written informed consent, participants were screened for eligibility. The criteria for inclusion consisted of male or female and aged between 18 and 60 years (inclusive) at the screening visit; have scar >1 year old and at least 10 cm^2^; adequate venous access in the left or right arm to allow collection of a number of blood samples; agreeance to highly effective methods of contraception during study period and 90 Days after last dose; able to understand, give consent, and comply with all scheduled study visits, procedures and restrictions.

Participants were not eligible to participate if they met any of the following exclusion criteria: clinically significant gastrointestinal, renal, hepatic, neurologic, hematologic, endocrine, oncologic, pulmonary, immunologic, psychiatric, skin, or cardiovascular disease or any other condition, that, in the opinion of the Investigator, would jeopardize the safety of the participant or impact the validity of the study results; current acute skin condition (*e.g.*, eczema, psoriasis, broken skin, wounds etc.) or large tattoos or excess hair at the study drug application site; pregnant or breastfeeding women; history of keloid scarring; history of immediate hypersensitivity to any medication or currently suffers from clinically significant systemic allergic disease; presence of a recent musculoskeletal injury or currently healing fracture; use of topical medications applied on treatment area; evidence of significant renal insufficiency, as indicated by an estimated creatinine clearance using the Cockcroft-Gault formula <80 mL/min at screening; receipt of blood, or loss or donation of 450 mL or more of blood within 90 Days or plasma donation within 14 Days before the first dose administration; have received an experimental therapy within 30 Days or 5 half-lives of the study drug, whichever is longer, prior to dosing; systemic infection other than common cold in the week prior to dosing. There were also some exclusion criteria in initial versions of the protocol which were later amended so that patients could be included at the Investigator’s discretion; if systolic blood pressure <90 or >140 mmHg, diastolic blood pressure <50 or >95 mmHg, and heart rate <40 or >100 bpm; if alanine aminotransferase (ALT), aspartate aminotransferase (AST), or bilirubin >1.5 × upper limit of normal; if haemoglobin, white blood cell (WBC), neutrophils, platelets < lower limit of normal. Tests may have been repeated once at the discretion of the Investigator.

### Study drug

The study drug was manufactured, packaged and labeled by Pharmaxis Ltd in accordance with Principles for Medicinal Products for use in human clinical Phase 1 trials in Australia. PXS-6302 is formulated as an oil in water cream which is obtained as a white to yellowish semi-solid. The cream is made by mixing the lipophilic phase (petroleum jelly 9.9 grs, paraffin oil 3.6 grs and cetostearyl alcohol 6.7 grs) with the aqueous phase (propylene glycol 38.3 g, water 36.9 g, NaH_2_PO_4_ 0.45 g, cetomacrogol 1000 1.7 g and 0.3-4% PXS-6302 (w/w), final pH ∼4.6-4.8) at 70°C, homogenizing and stirring until room temperature is achieved. A micro dosing device was provided containing sufficient cream for 35 days of once-per-day application. The participants were shown how to use the device during Day 1 visit and then provided with take-home instructions. The device allowed the application of 50 µL of cream per ‘click’. Participants used 4 ‘clicks’ to deliver 200 µL cream to the scar site of 10 cm^2^ and this was spread over the scar site using the dosing device that contained an application nozzle. Participants returned the device at Day 28 and Day 60 and were provided with a replacement at these visits. The study drug was allowed to be absorbed into the skin for at least 1 hour following application, after which there were no restrictions on touching or covering the area.

### Randomization

Cohort 1 was open label, and all participants were provided with PXS-6302 treatment and therefore no randomization occurred. Participants enrolled in Cohort 2 were randomized and placed in either the PXS-6302 treatment or placebo group (*n* = 21 per group). Participants were enrolled by the study coordinator, who assigned participants to intervention in accordance to start date. PXS-6302 and placebo treatment were randomized and blinded with a master code kept separate from staff involved in the trial. The allocation was conducted using permuted blocks to ensure balance between the treatment arms using blocks of 6.

### Procedure/trial visits

The study consisted of a screening period and a treatment period. Participants attended a screening visit between Day -28 and Day -2 to determine eligibility in the study. The screening visit included informed signed consent, taking a detailed medical and scar history, physical exam, measurement of vital signs and blood tests for clinical laboratory evaluations, including, haematology, clinical chemistry, and coagulation profiles. If the participant was eligible, participants were asked to attend a baseline visit (Day 1) within 28 Days. On Day 1, patients were randomized (Cohort 2 only) and prior to dosing, baseline assessments were performed, which included a physical exam, measurement of vital signs and scar assessments were performed, and a skin biopsy and blood samples were collected for baseline (pharmacodynamics and pharmacokinetics). The participant received treatment cream for at home dosing. The participants returned for assessments at Day 7, Day 21, Day 28, Day 60 and Day 90 in Cohort 1 and Day 28, Day 60 and Day 90 in Cohort 2.

### Data collection

Study data were collected and managed using REDCap (Research Electronic Data Capture) tools hosted by the Government of Western Australia, Department of Health (*19–20*). REDCap is a secure, web-based software platform designed to support data capture for research studies, providing an intuitive interface for validated data capture; audit trails for tracking data manipulation and export procedures; automated export procedures for seamless data downloads to common statistical packages; and procedures for data integration and interoperability with external sources.

### Primary outcomes-Safety

The primary outcome was to investigate the safety and tolerability of multiple applications over a 3-month period of PXS-6302 to an established scar. Assessments included: the incidence and severity of adverse events (AE); incidence of serious adverse events (SAE) and suspected unexpected serious adverse reactions; any clinically significant changes from baseline in clinical laboratory evaluations (haematology, clinical chemistry and coagulation blood results), vital signs and physical examinations.

### Secondary outcomes

#### Scar assessment

To assess if PXS-6302 improved scar appearance, scar assessments were conducted at baseline (Day 1) and at Day 28, Day 60 and Day 90. Scar assessment used POSAS Version 2.0 which includes two scales; Observer scale and a patient scale (*12*). Both scales contained six items that were scored numerically (from 1-10), with 1 reflecting normal skin and 10 indicating the worst imaginable scar. The observers rated scar vascularity, pigmentation, pliability, thickness, relief, and surface area. The patients rated pain, itching, color, pliability, thickness, and relief. The total score of both scales consisted of adding the scores of each of the six items (range, 6 to 60). The lowest score, 6, reflected normal skin, whereas the highest score, 60, reflected the worst imaginable scar. There was also an overall rating (1–10) for both the observer and patient and the investigator also asked the patient if they noticed any changes in the scar due to the treatment.

#### Optical coherence tomography imaging

Optical coherence tomography (OCT) imaging was performed on performed on a subset of consenting participants in Cohort 2 at the baseline (Day 1) and Day 90 *in vivo*, to assess the microvasculature and optical attenuation of the scar tissue. A 1,300-nm spectral-domain OCT scanner (upgraded TELESTO II, Thorlabs Inc., USA) was used for imaging with an axial and lateral imaging resolution of 5.5 µm (in air) and 13 µm, respectively (*21*). A custom imaging interface was used to mechanically couple the OCT probe with the skin to mitigate tissue motion during the acquisition of 3-D OCT scans (*11*). A 3-D scan was acquired from a tissue area of 6 × 4.5 mm within the selected 10 cm^2^ scar region for treatment with PXS-6302 or placebo cream, at both Day 1 and 90. Each 3-D scan comprised 1024 × 600 (× 2) × 1024 pixels in the lateral (*x* and *y*) and depth (*z*) directions, respectively, in which two repeated B-scans (*i.e.*, × 2) were acquired from each of the 600 *y* locations for subsequent OCT angiography (OCTA) processing to image the microvasculature. Care was taken to co-locate the same scar area for imaging at Day 1 and Day 90 using the skin features as landmarks.

Due to the large time span (*i.e.*, ∼90 days) of the two imaging sessions, a systematic change to the skin microvasculature can occur over different seasons which could confound the assessment of microvascular changes due to the treatment, although the imaging was performed at the same room temperature. To measure and subsequently correct for the systematic changes in microvasculature, an adjacent or contralateral 6 × 4.5-mm normal skin area outside the 10 cm^2^ treatment region was also selected for imaging at both Day 1 and Day 90, using the same scanning parameters as the treated scar region. In total, seven participants in the PXS-6302 group and another seven participants in the placebo group completed the two OCT imaging sessions.

#### Imaging and assessment of microvasculature

OCTA processing was subsequently applied to the acquired OCT scans to image and quantify the microvasculature (Fig. 4A). Each 3-D OCT scan was first processed by a speckle decorrelation algorithm, which was optimized for skin imaging. In particular, the decorrelation of the OCT intensity signal between each pair of repeated B-scans from the same lateral *y* locations was calculated, using a previously reported formula (*13*). This led to a decorrelation volume, in which blood vessels were highlighted by the high decorrelation values due to the high temporal changes in the OCT signal induced by the blood flow, in contrast to the static tissue with low decorrelation values. The decorrelation was further weighted by the corresponding original OCT intensity signal to reduce artificially high decorrelation caused by the OCT noise. A 2-D projection image of the vessels was then generated from the weighted decorrelation volume by taking the maximum decorrelation along the tissue depth direction from the skin surface to a depth of 500 µm, which was used to provide a qualitative assessment of the vessel network (Fig. 4B). The vessel projection image was then thresholded to calculate the total vessel area, which was divided by the total tissue area, resulting in the raw vessel area density.

To correct for the systematic change in microvasculature between Day 1 and Day 90, the raw vessel density of the scar tissue at Day 90 was further multiplied by a correction coefficient calculated based on the vessel densities of the untreated normal skin region as *VD_skin,D1_*/*VD_skin,D90_*. *VD_skin,D1_* and *VD_skin,D90_* are the raw vessel area densities of the untreated skin region at Day 1 and 90, respectively. The corrected vessel density of the treated scar region at Day 90 was then compared to the corresponding vessel density at Day 1 as a quantitative assessment of the microvascular changes caused by the treatment with PXS-6302 or placebo (Fig. 4C). During data processing, one patient from the PXS-6302 group showed high noise in the vessel image due to tissue movement during 3-D scan acquisition and was excluded from the analysis.

#### Imaging and assessment of optical attenuation

An attenuation imaging method (Fig. 4A) based on the single-scattering model was then applied to the same OCT scans to image and quantify the optical attenuation of scar tissue (*11*). OCT intensity signal in the original 3-D scans was first averaged between the two repeated B-scans from the same lateral locations to reduce OCT noise (Fig. 4D). Based on the single-scattering model, the averaged OCT A-scan signal (*i.e.*, 1-D depth signal at each lateral tissue location) comprised an exponential decay as well as two system factors, including the confocal function due to the focusing optics and the sensitivity roll-off due to the finite resolution of the detected spectrum. Prior to the estimation of the attenuation coefficient incorporated in the exponential decay component, the two system factors were first corrected using a calibration scan, acquired from a low-scattering solution of the polystyrene microspheres (Polybead®, Polysciences, Inc., Warrington, USA). The solution has a concentration of 5.69×10^9^ microspheres/ml with a uniform 0.5-µm diameter of the microspheres. Importantly, a 3-D correction was performed to particularly address the variation of the confocal function across the lateral locations within the 6 × 4.5-mm imaging field of view.

After correction, the OCT intensity signal was fitted to the exponential decay model to estimate the attenuation coefficient and corresponding goodness of fit, using weighted linear regression (*22*). In this study, an optimal length of the fitting window (200 µm) was used consistently for all scans, which was positioned in the top dermal region with reliable OCT signal. The fitting was performed to each averaged A-scan in the 3-D OCT scan, generating a 2-D *en face*attenuation coefficient image of the tissue for visualization (Fig. 4E). The histogram of the attenuation coefficients with a high goodness of fit was then used to show the distribution of the attenuation in the tissue and compared between Day 1 and 90 (Fig. 4E). The median attenuation was also calculated and compared between the Day 1 and 90 (Fig. 4F).

#### Human skin biopsy preparation

To investigate the pharmacodynamics of PXS-6302 when administered in multiple applications, skin punch biopsies (3 mm) were collected at Day 1 (pre-dose), Day 7, Day 28 and Day 90 for Cohort 1 and at Day 1 and Day 90 for Cohort 2. Samples were frozen at −80°C until analysis. To extract lysyl oxidases from the 3 mm human skin biopsy, epidermal and dermal layers of the skin were dissected, pre-cooled with liquid nitrogen and pulverized. Samples were washed thrice by homogenization with ice-cold wash buffer (0.15 M NaCl, 50 mM sodium borate, pH 8.0 with 0.25 mM phenylmethylsulfonyl fluoride and 1 µL/mL bovine aprotinin as protease inhibitors) at 100 µL/mg, centrifuged at 20,000 *g* for 10 minutes at 45°C and supernatant was used to measure PXS-6302 concentration. After the final washing step, the pellet was resuspended in extraction buffer (6 M urea, 50 mM sodium borate, pH 8.2 with 0.25 mM PMSF and 1 µL/mL bovine aprotinin) with ratio of buffer volume to tissue weight at 6:1. After 3-hour incubation at 4°C, the mixture was diluted with 8 × volume to tissue weight ratio of assay buffer containing 50 mM sodium borate (pH 8.2), centrifuged at 20,000 g for 20 minutes at 4°C. Lysyl oxidase concentration in the final supernatant was determined by direct ELISA assay and enzymatic activity in the skin extract was determined using a fluorescent based biochemical assay, as described below.

#### Fluorescent-based Lysyl oxidase activity assay

For the detection of Lysyl oxidase activity in the human skin biopsy extracts, the supernatant was spiked with pargyline hydrochloride and mofegiline hydrochloride at final concentrations of 0.5 mM and 1 µM, respectively, to inhibit other amine oxidases. The samples (25 µL) were then incubated with 600 µM β-aminopropionitrile (BAPN, a pan-lysyl oxidase inhibitor) or DMSO (control) for 30 minutes at 37°C and assayed by adding 25 µL of reaction mixture containing 50 mM sodium borate, 120 µM Amplex Red, 1.5 U/mL horseradish peroxidase, and 20 mM putrescine dihydrochloride as substrate, pH 8.2. Resorufin fluorescent signal intensity was measured every 2.5 min for 30 minutes at 45°C (with excitation and emission at 544 nm and 590 nm, respectively) and the enzymatic activity was indicated by the rate of the signal (kinetic value without BAPN). The percent activity from baseline was calculated by 100-100 (1-(signal over noise Day 90-1)/ signal over noise Day 1-1).

#### Protein and hydroxyproline measurement

Following lysyl oxidase extraction the remaining pellet underwent a reduction, hydrolysis and solid-phase extraction as previously described (*23–24*). An LC-MS/MS method was developed for determination of nine abundant amino acids (L-arginine, L-alanine, L-glycine, L-glutamic acid, L-serine, L-valine, L-proline, L-leucine and L-phenylalanine) concentration and hydroxyproline concentration in skin tissue. 1 μL of analyte after solid phase extraction was injected and separated on an Agilent Eclipse XDB-C18, 4.6 x 150mm, 5 µm column using isocratic elution. The mobile phase, 0.1% formic acid and methanol, was run at a flow rate of 0.5 mL/min with a total run time of 5 minutes. Amino acids and hydroxyproline were detected on a Thermo Scientific TSQ Endura Triple quadrupole by multiple reaction monitoring in positive electrospray ionization mode. The method was developed over a concentration range of 40–10,000 ng/mL. The total amino acid concentration was obtained by adding the selected amino acids. Hydroxyproline concentration was determined independently in the same run.

#### PXS-6302 measurement in plasma

To investigate the pharmacokinetics of PXS-6302 when administered in multiple applications over a 3-month period, blood samples (approximately 4 mL) were collected from participants in Cohort 1 at Day 1, Day 7, Day 21, Day 28, Day 60 and Day 90, and in Cohort 2 at Day 1 and Day 90 using K_2_-EDTA vacutainers. Blood samples were centrifuged to generate plasma and stored at −80°C until analysis. An LC-MS/MS method was developed and validated using a deuterated standard with a lower limit of detection of 0.01 ng/mL.

## Statistical analysis

Analyses presented were performed in the set of participants who had data available at each time-point. Twenty participants in the PXS-6302 treatment group completed the study (one participant lost to follow up). However, 2 participants had discontinued treatment prior to this point and therefore PD and PK analyses only included n=18 for all participants that completed treatment. All 21 evaluable participants from the placebo group completed the study (and treatment) and were included. Sensitivity analyses were performed a) including only completers and b) excluding data from participants with major deviations from the protocol and conclusions were consistent.

No formal analyses of safety data were performed. For POSAS, as the assumption of normality appeared adequate, changes from baseline to each time-point were analyzed using paired t-tests, and between group comparisons were performed using ANCOVA (adjusting for baseline, age of scar (<2 years/>=2 years) and type of scar (burn/non-burn) for POSAS endpoints).

Lysyl oxidase activity was calculated as a percentage of the baseline value. Other pharmacodynamic parameters (*e.g.*, hydroxyproline) were first normalized using the weight of the biopsy sample. The percentage change from baseline was then calculated for each patient. To estimate the size of the treatment effect, the difference in the means between treatment groups and standard error (SEM) were calculated; however non-parametric approaches were used to assess the significance of the differences (*i.e.*, Mann-Whitney tests) as these endpoints did not appear normally distributed.

## Data Availability

All data produced in the present study are available upon reasonable request to the authors

## Supplementary Materials

Data file S1

## Acknowledgments

The authors would like to thank Syntara and its drug discovery and clinical team for supporting this study. The authors would like to acknowledge Dr Kylie Sandy-Hodgetts for her contribution to setting up the study. The authors would also like to thank the clinical staff at Fiona Stanley Hospital Adult State Burns Service, in particular Sandeep B, Michal Nessler, Sarah Bache, Anna Goodwin-Walters and Joshua Wills for assistance with medical exams and with scar assessments. Thank you also to Grant Rowe and Aleksandra Miljevic from the Fiona Wood Foundation for assistance with processing biological samples. We would also like to thank all the participants who volunteered their time and effort for this study.

## Author contributions

Conceptualization: WJ, BC, FMW, MWF

Methodology: NM, PG, WJ, BC, AF, JL, FMW, MWF

Investigation: NM, PG, SR, HD, PHH, BC, BFK, FMW

Visualization: NM, PG, WJ, PHH, JL, AF, MWF

Funding acquisition: WJ, BC, FMW

Project administration: NM, WJ, AF, MWF

Supervision: WJ, BFK, BC, FMW, MWF

Writing – original draft: NM, PG, WJ, AF, MWF

Writing – review & editing: NM, PG, SR, HD, BC, BFK, WJ, AF, FMW, MWF

## Data and materials availability

All data are available in the main text or the supplementary materials.

## References and Notes

1. PPM van Zuijlen, JJB Ruurda, HA van Veen, J van Marle, AJM van Trier, F Groenevelt, RW Kreis, E Middelkoop. Collagen morphology in human skin and scar tissue: no adaptations in response to mechanical loading at joints Burns 29,423–431(2003)

2. L Maknuna, H Kim, Y Lee, Y Choi, H Kim, M Yi, HW Kang. Automated Structural Analysis and Quantitative Characterization of Scar Tissue Using Machine Learning. Diagnostics 12, 534 (2022).

3. M.G Jeschke., F. M. Wood, E. Middelkoop, A. Bayat, L. Teot, R Ogawa and G.G Gauglitz. Scars. Nat Rev Dis Primers 9, 64 (2023).

4. L. I. Smith-Mungo, H. M. Kagan. Lysyl oxidase: properties, regulation and multiple functions in biology. Matrix Biol 16, 387–398 (1998).

5. H. M. Kagan, W. Li. Lysyl oxidase: properties, specificity, and biological roles inside and outside of the cell. J Cell Biochem 88, 660–672 (2003).

6. D. L. Clarke, A. M. Carruthers, T. Mustelin, L. A. Murray, Matrix regulation of idiopathic pulmonary fibrosis: the role of enzymes. Fibrogenesis Tissue Repair 6, 20 (2013).

7. R. Fleischmajer, L. Jacobs, E. Schwartz, L. Y. Sakai, Extracellular microfibrils are increased in localized and systemic scleroderma skin. Lab Invest 64, 791–798 (1991).

8. P. Sivakumar, C. Kitson, G. Jarai, Modeling and measuring extracellular matrix alterations in fibrosis: challenges and perspectives for antifibrotic drug discovery. Connect Tissue Res 60, 62–70 (2019).

9. A. J. van der Slot, A. M. Zuurmond, A. J. van den Bogaerdt, M. M. Ulrich, E. Middelkoop, W. Boers, H. Karel Ronday, J. DeGroot, T. W. Huizinga, R. A. Bank, Increased formation of pyridinoline cross-links due to higher telopeptide lysyl hydroxylase levels is a general fibrotic phenomenon. Matrix Biol 23, 251–257 (2004).

10. N. Chaudhari, A. D. Findlay, A. W. Stevenson, T. D. Clemons, Y. Yao, A. Joshi, S. Sayyar, G. Wallace, S. Rea, P. Toshniwal, Z. Deng, P. E. Melton, N. Hortin, K. S. Iyer, W. Jarolimek, F. M. Wood, M. W. Fear, Topical application of an irreversible small molecule inhibitor of lysyl oxidases ameliorates skin scarring and fibrosis. Nat Commun 13, 5555 (2022)

11. P Gong, RA McLaughlin, YM Liew, PR Munro, FM Wood, DD Sampson. Assessment of human burn scars with optical coherence tomography by imaging the attenuation coefficient of tissue after vascular masking. J Biomed Opt. 19: :21111.(2014)

12. A. L. van de Kar, L. U. Corion, M. J. Smeulders, L. J. Draaijers, C. M. van der Horst, P. P. van Zuijlen, Reliable and feasible evaluation of linear scars by the Patient and Observer Scar Assessment Scale. Plast Reconstr Surg 116, 514–522 (2005).

13. P Gong, S Es’haghian, K.A Harms, A. Murray, S. Rea, B. F. Kennedy, F.M. Wood, D.D Sampson, and R. A. Mclaughlin. Optical coherence tomography for longitudinal monitoring of vasculature in scars treated with laser fractionation. J Biophotonics. 9 :626–636. (2016)

14. A. Hauge., E.K. Rofstad. Antifibrotic therapy to normalize the tumor microenvironment. J Transl Med 18, 207 (2020).

15. PW Hu, CK Chen, YH Hsiao, CY Weng, YC Lee, KC Su, JY Feng, KT Chou, DW Perng , HK Ko. Correlations between blood vessel distribution, lung function and structural change in idiopathic pulmonary fibrosis. Respirology. 29: :962–968 (2024).

16. C Polydorou, F Mpekris, P Papageorgis, C Voutouri, T. Stylianopoulos. Pirfenidone normalizes the tumor microenvironment to improve chemotherapy. Oncotarget. 20. 24506–24517. (2017)

17. P. R. Burchard, L.I. Ruffolo LI, N.A. Ullman, B.S. Dale, Y.A. Dave, B.K. Hilty, J Ye, M. Georger, R. Jewell, C. Miller, L. De Las Casas, W. Jarolimek, L. Perryman, M.M. Byrne, A. Loria, C. Marin, M. Chávez Villa, J.J. Yeh, B.A. Belt, D.C. Linehan, R. Hernandez-Alejandro. Pan-lysyl oxidase inhibition disrupts fibroinflammatory tumor stroma, rendering cholangiocarcinoma susceptible to chemotherapy. Hepatol Commun. 8:e0502. (2024)

18. J.L. Chitty, M. Yam, L. Perryman, A. L. Parker, J. N. Skhinas, Y. F. I. Setargew, E. T. Y. Mok, E. Tran, R. D. Grant, S. L. Latham, B. A. Pereira, S. C. Ritchie, K. J. Murphy, M. Trpceski, A. D. Findlay, P. Melenec, E. C. Filipe, A. Nadalini, S. Velayuthar, G. Major, K. Wyllie, M. Papanicolaou, S. Ratnaseelan, P. A. Phillips, G. Sharbeen, J. Youkhana, A. Russo, A. Blackwell, J. F. Hastings, M. C. Lucas, C. R. Chambers, D. A. Reed, J. Stoehr, C. Vennin, R. Pidsley, A. Zaratzian, A. M. Da Silva, M. Tayao, B. Charlton, D. Herrmann, . Nobis, S. J. Clark, A. V. Biankin, A. L. Johns, D. R. Croucher, A. Nagrial, A. J. Gill, S. M. Grimmond, Australian Pancreatic Cancer Genome Initiative (APGI), Australian Pancreatic Cancer Matrix Atlas (APMA), M. Pajic, P. Timpson, W Jarolimek & T. R. Cox. A first-in-class pan-lysyl oxidase inhibitor impairs stromal remodeling and enhances gemcitabine response and survival in pancreatic cancer. Nat Cancer 4, 1326–1344 (2023).

19. P. A. Harris, R. Taylor, B. L. Minor, V. Elliott, M. Fernandez, L. O’Neal, L. McLeod, G. Delacqua, F. Delacqua, J. Kirby, S. N. Duda, R. E. Consortium, The REDCap consortium: Building an international community of software platform partners. J Biomed Inform 95, 103208 (2019).

20. P. A. Harris, R. Taylor, R. Thielke, J. Payne, N. Gonzalez, J. G. Conde, Research electronic data capture (REDCap)--a metadata-driven methodology and workflow process for providing translational research informatics support. J Biomed Inform 42, 377–381 (2009)

21. Q. Wang, P. Gong, B. Cense, and D. D. Sampson, Short-time series optical coherence tomography angiography and its application to cutaneous microvasculature. Biomed. Opt. Express 10, 293–307 (2019)

22. P. Gong, M. Almasian, G. van Soest, D. de Bruin, T. van Leeuwen, D. Sampson, & D. Faber. Parametric imaging of attenuation by optical coherence tomography: Review of models, methods, and clinical translation,” Journal of Biomedical Optics 25, 040901 (2020)

23. A. Findlay, C. Turner, H. Schilter, M. Deodhar, W. Zhou, L. Perryman, J. Foot, A. Zahoor, Y. Yao, R. Hamilton, M. Brock, C. Raso, J. Stolp, M. Galati, D. Hamprecht, B. Charlton, W. Jarolimek, An activity-based bioprobe differentiates a novel small molecule inhibitor from a LOXL2 antibody and provides renewed promise for anti-fibrotic therapeutic strategies. Clin Transl Med 11, e572 (2021).

24. A. Joshi, A. Zahoor, A. Buson, Measurement of Collagen Cross-Links from Tissue Samples by Mass Spectrometry. Methods Mol Biol 1944, 79–93 (2019).

